# Development of a customised data management system for a COVID-19-adapted colorectal cancer pathway

**DOI:** 10.1101/2020.10.05.20206920

**Authors:** Frances Gunn, Janice Miller, Malcolm G Dunlop, Farhat V N Din, Yasuko Maeda

## Abstract

**Purpose:** The COVID-19 pandemic posed an unprecedented challenge to healthcare systems around the world. To mitigate the risks of those referred with possible colorectal cancer during the pandemic we implemented a clinical pathway which required a customised data management system for robust operation. Here, we describe the principal concepts and evaluation of the performance of a spreadsheet-based data management system.

**Methods:** A system was developed using Microsoft Excel^®^ 2007 aiming to retain the spreadsheets inherent intuitiveness of direct data entry. Data was itemised limiting entry errors. Visual Basic for Applications (VBA) was used to construct a user-friendly interface to enhance efficiency of data entry and segregate the data required for operational tasks. This was done with built-in loop-back data entry. Finally data derivation and analysis was performed to facilitate pathway monitoring.

**Results:** For a pathway which required rapid implementation and development of a customised data management system, the use of a spreadsheet was advantageous due to its user-friendly direct data entry capability. Its function was enhanced by UserForm and large data handling by data segregation using VBA macros. Data validation and conditional formatting minimised data entry errors. Computation by the COUNT function facilitated live data monitoring on a dashboard. During the three months the pathway ran for, the system processed 36 nodal data points for each of the included 837 patients. Data monitoring confirmed its accuracy.

**Conclusion:** Large volume data management using a spreadsheet system is possible with appropriate data definition and VBA programmed data segregation. Clinicians’ regular input and optimisation made the system adaptable for rapid implementation.

## Introduction

The COVID19 pandemic has brought about immense challenges to healthcare systems across the world. Care was directed towards those most critically ill whilst other hospital services were curtailed or stopped completely, including endoscopy – the gold standard test for diagnosing colorectal cancer.

We introduced a COVID-adapted colorectal cancer pathway in an attempt to mitigate the risks of delayed and missed colorectal cancer diagnosis using a combination of the available tests (quantitative faecal immunohistochemistry test [qFIT] and CT modified preparation scan) [1]. Operation of any clinical pathway requires a sound information management system in order to minimise errors, process the flow of patients, and monitor outcomes. Real-time feedback is important to analyse performance (in this case cancer detection) and facilitate support from other clinicians and management [2]. Best practice is to have such a pathway embedded in an existing health informatic system [3]. However, it was challenging to facilitate such a rapid change in practice in a short time when the healthcare service was adapting to deal with the COVID-19 pandemic.

Here, we report our experience of developing a bespoke data management system that was utilised to ensure risk mitigation in patients who were presenting with symptoms suspicious for colorectal cancer during this time. We describe the local set up and planning of a database system using Microsoft Excel^®^ and Visual Basic for Application (VBA).

## Methods

### Considerations made for the design of the data management system

The pathway involved several different investigations with patients being signposted to the appropriate tests in a stepwise fashion depending on referral information and initial test results. This was done to maximise the use of available testing and ensure those patients deemed to be at highest risk were prioritised. In addition to clinical flow, several letters were created to explain the rationale and steps of the pathway to patients and general practitioners along with safety netting advice as shown in figure 1.

During the design of the data management system the requirement of both administrative and clinical information was considered to estimate the information flow and risks associated with data handling. This was done taking into account the availability of resources and volume of clinical practice. Several questions were therefore asked to estimate the quality and quantity of information flow including the amount of patients estimated to be entered into the pathway, how much nodal information and classification were required to signpost patients into the correct pathway arm according to test results, what other data items were needed to make live data presentation possible, particularly relating to cardinal outcomes and monitoring of pathway performance (e.g. number of patients, cancer detection rate) and finally what information was required to manage administrative tasks (e.g. sending out different types of letters correctly or monitoring of when tasks were completed)?

### Risk assessment

A risk assessment was undertaken considering the pros and cons of different information methods, namely paper based vs spreadsheet vs database vs use of the existing hospital electronic system. Consideration was made to the time, resources and expertise required to build a new, robust and safe data management system in a short space of time. We also looked at the number of staff available to operate the pathway, their level of IT skills and amount of induction and teaching required to introduce it. Finally, we considered the available support and feedback from clinical and management teams.

### Estimation of information volume

The volume of information required for the pathway was estimated from proxy indicators based on historical data, such as the number of cancers diagnosed (>500 per year), the number of urgent referrals (>3000 per year) and the availability of emergency diagnostic tools during the pandemic (almost null). We estimated the number of patients to be referred to the pathway would be approximately 500 in two months. We also factored in that the workload may continuously increase due to uncertainties surrounding the pandemics duration and the prospect that services would not return to full capacity for the foreseeable future.

Given the multiple tests built into the pathway and the need to stratify patients according to their qFIT result, the volume of information expected was deemed beyond the capabilities of a paper based system. It was felt this approach would lack connectivity to live results, be insensitive to changes in patients status and be difficult to filter relevant information from multiple records thus reducing efficiency.

### Key data items and hierarchical order

Key data items required for pathway management were enlisted and defined clearly. Entry to the pathway or not (to filter out requests of tests out with the pathway) was the cardinal item in addition to the urgency of the referral category. There were two personal identifications (ID) for each patient (CHI: community health index number and UHPI: system allocated unique reference number) in the existing electronic hospital record system. The UHPI was designated as the primary key ID due to the absence of CHI in some patients (e.g. visitor from another country or area of the United Kingdom or those patients not registered with a General Practitioner). Each UHPI contains a 9-digit number that ends with a letter compared with a 12-digit numerical CHI. It therefore provided a better property as string information and linkage to electronic health records.

The type, combination and results of investigations were pathway specific key turning points for signposting clinicians to the next step for their patients and were recorded as nodal data type so that they could be grouped into lists or put in hierarchical order when required. Any information was further broken down into elemental data and classified into categories (e.g. yes or no, 0 or 1).

### Data management system using spreadsheets

It was not possible to introduce customised change into the existing hospital electronic record system or rapidly acquire database software. We therefore explored the option of using a spreadsheet. Although spreadsheets such as Microsoft Excel^®^ or Google Sheets are some of the commonly used tabular data management tools in clinical practice and research, they are known to be inefficient for handling large volumes of data [4]. However, a spreadsheet has a user-friendly interface which favoured our situation where there was little time for induction and training of staff due to the pressing need to swiftly introduce the pathway. Spreadsheets also allow computation with capabilities for ad-hoc analysis. The software (Microsoft Excel^®^ (2007)) had already been universally installed and was available on all hospital system hard disks. Files could be maintained on a shared drive within the secure hospital system.

### Practical solutions to overcome the limitations of spreadsheets and concepts of the data management system

We developed a system to maximise the functionality of the spreadsheet and incorporate VBA whilst minimising its limitations by segmentation of data based on the following concepts: We kept the master spreadsheet (parent) as a .xlsx file and limited the use of VBA to child files. We retained the spreadsheet’s inherent intuitiveness of direct data entry. Data was Itemised and drop down menus used thus taking advantage of the Data Validation function to limit entry errors. A parallel option of UserForm constructed by VBA as a user-friendly interface, to enhance efficiency of data entry and provide overview of results was used. Formulae were utilised to allow derived computation and facilitate live monitoring of the pathway activities. Data were segregated for operational tasks with built-in loop-back data entry. The flow of information was controlled via multiple programmed .xlsm with one-way data traffic to avoid overwriting existing data in the parent file.

## Results

A master spreadsheet was constructed which consisted of 36-items including demographical information (e.g. ID, age, sex) and specific data for the pathway operation (tests, results and tasks). Information from referral letters, blood results, and past medical history (56 items) were collected as general background information.

### Maintaining the master file and minimising data entry errors

The master spreadsheet was saved and kept as an .xlsx file to prevent receiving malicious VBA codes. Direct data entry was possible for all 36 data items. There were several mechanisms introduced to minimise errors from intuitive data entry.

A spreadsheet was created using the formula:

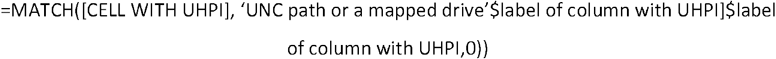

to find the matching CHI/UHPI in the main spreadsheet in order to prevent duplicate entry to the master file when a new request for a test came in (figure 2). This was used in addition to CHI and UHPI columns with ‘Conditional Formatting’ highlighting any duplicate cell.

Entry using free text was limited to two data items. These were to include useful information such as comments from patients during telephone contact or tailored information to be added to patient letters e.g .findings from CT scan results. All other information was divided into elemental items that could be recorded in categorical entries using Microsoft Excel’s Data Validation and drop down list function. For entry of dates, Data Validation was used to restrict entries to a certain time period to avoid gross error (e.g. wrong year). Furthermore, cell format and Text to Columns function was used periodically to apply the correct format.

In addition, any item that could be linked to unique IDs (CHI, UHPI) were automated by using the formula. Patient’s sex could be worked out from the second to the last digit of the CHI; even numbers corresponding to females and odds to males. The following formula was used to avoid errors once the CHI was copied in from the electronic health record:

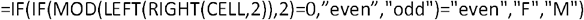

IF function was also used in a formula =IF(CELL=““,”“,”to be ordered”) during the processing of test results. As soon as the decision was made regarding the next step in the pathway, ‘to be ordered’ was shown in the order status until a physical request had been placed on the hospital electronic record. The user physically had to click on a drop down to change it to ‘ordered’ and until this occurred it remained on the ‘outstanding tasks’ list which was created by VBA (described in detail later).

### Development of the data entry form

A UserForm was created for efficient data entry which was directly linked to the master spreadsheet. It consisted of a function to extract data for display, update information of existing patents and add new patients with triaging information (Figure 3). This was based on VBA to simply display the patient’s information and results of tests. The ‘update’ button allowed the user to overwrite information in a row with the matching UHPI in the master spreadsheet. The button ‘new patient’ was programmed to enter data in the next row after the last existing row. Data entries were restricted to entering IDs and triaging information thus completely segregating two sections of data entries. Entry date was regulated by the use of VBA property CDATE.

### Data segregation

Furthermore, VBA was used to filter data according to different purposes. Appropriate segregation of information was required to outline tasks and streamline the daily operation of the complex pathway taking care to avoid fragmentation of information.

A child file (‘all_patients_on_pathway.xlsm’) was created using VBA macros to filter all patients included in the pathway from the master file using a key indicator which was nodal data [e.g. whether the pathway was included (yes or no)]. The function of Advanced Filter VBA was used due to its speed and function to filter by column values with the use of AND or OR logic. The filtered data of all patients on the pathway was then used as the new ‘parent’ file which was updated daily to source most of the tasks and reference lists relating to the pathway operation. As the data in this file was updated only once per day, it could be up to 24 hours old. However, this did not affect extracting the task lists, for example, to chase patients who had not returned a qFIT test for more than 6 weeks (=“<“&(TODAY()-42). In a similar manner, several task lists were programmed utilising elemental data.

### Overall framework of the data management system

Flow of information via multiple programmed .xlsm was mostly one-way traffic to avoid overwriting existing data in the master file. Direct access to the master file was restricted to a super-user file such as ‘tracker.xlsm’ to monitor pathway performance. The file filtered patients on the pathway in a similar manner as ‘all_patients_on_pathway.xlsm’ and had additional built-in calculations to monitor the pathway operation. Other task files that needed feedback of results or information on task completion were limited to three.

In the ‘tracker.xlsm’, calculations were performed using a number of formulae, mainly the COUNTIF and COUNTIFS function. For example, the number of patients who had qFIT as the first test could be counted by a simple COUNTIF with counting ‘qFIT’ (=COUNTIF(Results!J:J,”qFIT only”) whilst further breakdown according to result group were done by COUNTIFS (eg for results 10-79, =COUNTIFS(Results!E:E,”pathway”, Results! Results!J:J,”qFIT only”, Results!H:H,”>9”, Results!H:H,”<80”). These built-in formulae allowed monitoring of both performance and figures as a one-screen dashboard in real-time.

In addition, the COUNT function was used to make a user-friendly pathway tree. There were multiple formulae to count the direction of patients’ flow which helped the ground staff to correctly signpost the patient to the next step or send out the appropriate letters.

An overview of the data management and one way flow systems are summarised in figures 4 and 5. Patient flow is summarised in figure 6, the pathway dashboard in figure 7 and pathway tree in figure 8.

### Assessment of the quality of data from the pathway

Between 1^st^ April and 30^th^ June, 837 patients were added to the pathway. Entered patients were randomly sampled and manually cross-checked with their hospital electronic record to assess the accuracy of data entries. All duplicate entries were picked up by either ‘Conditional Formatting’ or ‘Row number finder’. Failed entry of results (cell value=NULL) were picked up by task lists such as chasing qFIT/CT or phone calls to patients and were mostly due to patients not submitting tests or failing to attend appointments. Any incomplete test orders were detected by ‘outstanding tasks’ which was sourced by advanced filtering by selecting ‘to be ordered’ or cells with NULL values (=<>).

## Discussion

Development of the current data management system was born out of the need to rapidly set up a pathway for patients referred with symptoms of colorectal cancer during the COVID-19 pandemic. Modification of any existing hospital electronic systems is challenging at the best of times as healthcare industries tend to have multiple but disparate information systems. Maintenance and modification of any system is often outsourced to external providers and is difficult to reflect the acute need required in clinical practice. During the pandemic when the rapid launch of a risk mitigation strategy was needed, the availability of expertise to build a bespoke database was severely limited. The concept and design of this system could be utilised in any resource constrained setting when the rapid development and introduction of a data management system is required.

Times of crisis are tests of resilience, however, historically they offer opportunities for new ideas and innovation. The development of the current system presented a unique opportunity for clinician centred design. It required understanding of both the clinical pathway and the hospital IT-systems. Clinicians’ continuous input reflecting the pathway practicalities and operation allowed identification of workflow bottlenecks and solved any issues promptly. When information processing was unclear, staff were supported by senior medics involved in decision making which allowed fine tuning of the system without delay.

Collection of accurate information is key to avoid error and facilitate robust pathway operations. Considerable efforts and attention to detail were required to apply multiple manual checks to run day to day tasks safely whilst the pathway evolved. Data cleaning to ensure all correct codes were applied required time and effort but this was expected due to the makeshift nature of the system and the inherent limitations of a spreadsheet being able to function as a bona fide database. From a governance perspective, this data validation process was heavily dependent on personnel. However, periodic discussion within the team and monitoring of the pathway helped to ensure accuracy and flow of the patients. Presentation of real-time data and feedback from other clinicians was useful to develop the pathway and make necessary adjustments to data definition and validation. The flexibility and user-friendliness of the spreadsheet allowed staff to familiarise themselves with the system in a relatively short period of time without much training. The system was easily adaptable as the pathway evolved with the rapid changes within the healthcare system.

Although dealing with a pandemic is a situation unique to most, managing large volumes of data using a spreadsheet is quite common both in clinical practice as well as research. The described concept is applicable to any data management system construction requiring speed and flexibility in a resource limited situation.

## Conclusion

Large volume data management using a spreadsheet system is possible with appropriate data definition and Visual Basic for Application macro programmed data segregation. This can increase efficiency of the system and circumvent some of the limitations of spreadsheet function. Clinicians input and continuous optimisation made the system adaptable and suitable for bespoke data management in a resource-limited setting.

## Supporting information

Images

## Data Availability

Availability of data and material: A data cleaned file is available as a reference
Code availability: Available if contacted

## Declarations

### Acknowledgements

The authors would like to thank Neil Gunn for their review of technical details.

### Availability of data and material

A data cleaned file is available as a reference

### Code availability

Available if contacted

### Authors’ contributions

Yasuko Maeda designed and ran the spreadsheet. Yasuko Maeda, Janice Miller and Frances Gunn developed and refined the system and assessed its practical application. Yasuko Maeda, Malcolm Dunlop and Farhat Din contributed to performance analysis and data interpretation. Yasuko Maeda and Janice Miller wrote the manuscript. All authors critically appraised and approved the final version.

